# Neural networks for inhibitory function and error detection in younger and older adults for early detection of cognitive decline: a comparative study

**DOI:** 10.1101/2025.11.20.25340678

**Authors:** Kazumasa Ukai, Kazuhei Nishimoto, Hiroki Ito, Ryosuke Yamauchi, Kouta Maeda, Osamu Katayama, Shin Murata, Kiichiro Morita, Takayuki Kodama

**Author notes:** Corresponding Author: Kazumasa Ukai, Graduate School of Health Sciences, Kyoto Tachibana University 34 Yamada-cho, Oyake, Yamashina-ku, Kyoto 607-8175, Japan.

## Abstract

Cognitive function can decline irreversibly with age, potentially progressing to dementia. Intervention during the preclinical stage is considered effective, but evaluation often requires large-scale equipment and tests. A decline in inhibitory function precedes and is involved in a general decline in higher-order brain functions, making its assessment a key target for early detection. We focused on error-related negativity (ERN) to capture the neural network of inhibitory function. While larger ERN amplitudes indicate higher error detection ability and correlate with inhibitory function, the causal relationship, mutual influences, and age-related effects in older adults remain unclear.

This study measured brain activity during an inhibitory task in young and older adults to examine the neural networks related to error detection and inhibitory function. We used LORETA iCoh Full Vector Field analysis to verify directional connectivity.

In the elderly group, during error responses, we observed significantly stronger beta-band directionality from the ventral anterior cingulate cortex (ACC) to the left frontal pole. Between the ventral and dorsal ACC, we also found significantly stronger directionality in the theta, alpha, and beta bands. During correct responses, they showed significantly stronger alpha and beta-band directionality from the left dorsolateral prefrontal cortex (DLPFC) to the right frontal pole.

Compared to the elderly, the young group exhibited significantly stronger mutual directionality between the ventral and dorsal prefrontal cortices in the alpha and beta bands. They also showed widespread, significantly strong directionality among the ACC, bilateral DLPFC, and frontal poles.

These results suggest that error detection ability is important for the normal functioning of inhibitory control in older adults. Furthermore, an age-related decline in metacognitive abilities may be associated with impaired inhibitory function. The insights from this study can contribute to developing risk prediction models for cognitive decline and establishing effective preventive strategies.

## 1. Introduction

Age-related decline in higher brain function is a phenomenon caused by structural changes in the brain. Age-related structural changes in the brain are caused by the accumulation of subtle changes, such as a decrease in the number of neurons, reduction in synapses, and changes in neurotransmitters [1]. These changes are thought to lead to a decrease in brain plasticity and efficiency of neural networks, resulting in impaired information processing abilities [2]. Recent diffusion tensor imaging studies have demonstrated that subtle structural changes in the white matter are closely associated with cognitive decline [3], particularly with reduced connectivity in the prefrontal–parietal network, which is associated with impaired executive function [4]. These structural changes progress with age, becoming particularly prominent from the 50s onward, as revealed by longitudinal studies [5]. Higher educational attainment and rich intellectual activity enhance cognitive reserves, thereby mitigating the effects of age-related structural changes in the brain [6]. The concept of cognitive reserve has gained attention as a theoretical framework for explaining why individuals with similar structural changes in the brain may exhibit different degrees of cognitive decline [7]. Particularly, higher educational attainment and continued intellectual activity enhance cognitive reserve and improve functional resilience against structural changes in the brain [8]. These findings highlight the importance of early preventive interventions, suggesting that age-related declines in higher brain function are not necessarily inevitable and that their progression can be delayed or mitigated through appropriate preventive measures [9].

A detailed examination of the process of age-related changes in higher brain functions reveals that their progression is gradual [10]. The first stage is the preclinical stage of dementia, in which mild impairment of higher brain function occurs without significant impairment in daily life or clear declines in objective cognitive function tests [11]. From this stage, the condition progresses gradually, passing through a stage of mild cognitive impairment (MCI), in which mild functional decline can be detected through objective cognitive function tests, ultimately leading to dementia [12]. Recent longitudinal studies have reported that changes in functional connectivity within brain networks occur during the preclinical stages of dementia [13]. In particular, changes in the interaction between the default mode network and the executive control network have been suggested as potential early markers of cognitive decline [14]. Additionally, machine learning analyses have suggested the potential of predicting the risk of cognitive decline using brainwave data [15]. Active interventions, such as cognitive rehabilitation, during the preclinical stage of dementia, when no clinical symptoms are present, can delay the decline of higher brain functions or maintain the current state to some extent [16]. However, to accurately assess higher brain function in the preclinical stage, advanced medical devices and specialized examinations, such as functional magnetic resonance imaging and biomarkers, are required [17]. Therefore, data on changes in higher brain function in the preclinical stages of dementia are currently limited. In addition, regarding age-related changes in neural substrates, the mechanisms underlying the decline in individual functions and how they interact with each other remain poorly understood [18].

In human cognitive function, inhibitory function is a key component of executive function [19]. This function enables appropriate actions aligned with goals by suppressing inappropriate responses or behaviors [20]. Recent meta-analyses have revealed that the right inferior frontal gyrus, pre-supplementary motor area, and ACC play central roles in inhibitory function [21]. Additionally, the functional connectivity between these regions correlates with inhibitory function performance [22]. Inhibitory function has multiple aspects, including response, cognitive, and interference inhibitions, each interacting with different cognitive processes [23]. Response inhibition is involved in stopping ongoing actions, cognitive inhibition suppresses unnecessary thoughts or memories, and interference inhibition controls the influence of task-irrelevant information [24]. These functions form the basis of adaptive behaviors in daily life and play important roles in learning and social interactions [25]. In particular, inhibitory function shows a relatively early decline with age [26] and has a significant impact on overall cognitive function, making it important to examine its functional mechanisms in relation to other cognitive functions. Inhibitory functions are closely related to various cognitive functions, particularly working memory, attention control, and metacognitive abilities [27]. These functions share the prefrontal cortex and mutually influence each other [28]. For example, working memory is responsible for maintaining and updating goal-relevant information, whereas inhibitory function suppresses inappropriate responses based on that information [29]. Attention control enables the maintenance of attention to relevant information and the shifting of attention away from irrelevant information, thereby supporting efficient functioning of the inhibitory function [30].

Error detection functions identify errors that occur during behavioral or cognitive processes and facilitate action correction based on these errors [31]. Error detection plays a crucial role in behavioral adjustment and serves as the foundation for adaptive behavior [32]. The processing of error detection involves both metacognitive conscious processing and distinct unconscious processing mechanisms [33]. Metacognitive error detection primarily occurs in the frontal polar cortex (FPC) [34] and involves conscious evaluation of the outcomes of actions or thoughts and making necessary corrections. This process involves explicit awareness of one’s cognitive state or the outcomes of actions, enabling more complex cognitive control [35]. This function plays a particularly important role in problem-solving and learning situations and supports long-term behavioral adjustment [36]. In contrast, unconscious error detection has a different neural basis from conscious error detection and is primarily associated with the ACC [37]. This processing mechanism rapidly detects potential errors before the results of the actions are consciously recognized, enabling immediate behavioral corrections [38]. This unconscious error detection process is observed as the error-related negativity (ERN), one of the event-related potential components [39]. ERN is a negative potential that appears 50–150 msec after the response, and its amplitude is correlated with the sensitivity of error detection [40]. These two distinct processing processes operate at different stages—immediate processing and deliberative processing—while leveraging their respective characteristics to contribute to behavioral control [41].

The interrelationships among the three systems—inhibitory function, metacognitive error detection, and unconscious error detection—are not yet fully understood. These processes are neurologically processed in different brain regions, but further investigation is required to understand how they coordinate, particularly regarding the order and interactions of these processes [42]. For example, the detailed mechanisms by which initial error detection signals in the ACC influence processing in the FPC and how this ultimately contributes to the adjustment of inhibitory functions remain unclear [43]. Additionally, empirical investigations are required to clarify how these processing mechanisms are involved in a stepwise manner, from immediate response control to long-term behavioral adjustment [44]. Recent studies have suggested that the dynamic interactions between these functions may enable flexible cognitive control [45]. Elucidating such relationships can contribute to a better understanding of cognitive dysfunction and the development of effective intervention methods, making this an important research topic.

This study aimed to clarify how conscious and unconscious error detection abilities are involved in the decline in inhibitory functions with age. Clarifying the relationship between conscious and unconscious error detection abilities and decline in inhibitory functions may provide important insights into the mechanisms underlying age-related decline in inhibitory functions. This is expected to lead to a new understanding that adds the functional aspect of error detection to the previously identified neural basis of prefrontal cortex dysfunction. Furthermore, this study aimed to clarify the dynamic interactions of neural networks involved in cognitive function using the latest brainwave analysis technique, Low-Resolution Electromagnetic Tomography (LORETA) iCoh Full Vector Field analysis, to examine the directed connections between brain regions in detail.

## 2. Participants and Methods

### 2.1 Participants

This study comprised 17 elderly men (n = 9) and women (n = 8) residing in the community (mean age, 78.3 ± 4.2 years) and 15 healthy adult men (mean age, 20.3 ± 0.5 years) as the control group. The inclusion criteria were a sleep duration of ≥ 6.5 h on the previous day and no caffeine intake for 6 h prior to the experiment. The exclusion criteria included a Mini-Mental State Examination score of ≤ 27 [46], visual impairments that would interfere with task performance, motor or sensory impairments that would interfere with task performance, orthopedic disorders or disorders related to higher brain functions, and medication that could affect brain activity. Participant recruitment for this study started on 2 October 2023 and ended on 31 March 2024. The research content was explained verbally and in writing, and written informed consent was obtained. This study was approved by the Kyoto Tachibana University Research Ethics Committee (approval number: 23-45) and was conducted in accordance with the Declaration of Helsinki.

### 2.2 Experimental procedures

In this study, after a pre-assessment, brainwave activity was measured during a 2-min resting state (pre-Rest) while seated in a chair. During this period, the participants were instructed to maintain a stable posture and avoid unnecessary body movements. To maintain a more natural state of wakefulness, measurements were taken with the eyes open. After completion, the participants were presented with multiple target stimulus conditions (geometric stimuli) on a digital display in front of them and were instructed to press or not press a switch button as quickly as possible in response to the stimuli. This task was referred to as the “inhibition task” in this study. During the inhibition task, the participants were instructed to focus their gaze on the center of the monitor and perform the task. When the presented stimulus was a figure with five arrows arranged horizontally in the same direction (match stimulus), the participants were instructed to “not press the button.” When the presented stimulus was a figure with five arrows arranged horizontally in the same direction with only the middle arrow pointing in the opposite direction (mismatched stimulus), the participants were instructed to “press the button.” The number of stimuli was set to 30 trials for each side for left-right matched stimuli and 20 trials for each side for left-right mismatched stimuli, for a total of 100 trials. Each stimulus was presented for 300 ± 0.5 msec. To prevent anticipatory responses from the participants, stimuli were presented randomly at intervals of 1995 ± 99 msec. A multitrigger system (Medical Try System Co., Ltd.) was used as the stimulus device (Figure 1). After completing the task, the participants were seated again with their eyes open, and brain wave activity was measured during a 2-min rest period (post-rest). Prior to the post-rest measurement, short breaks were provided, as needed, to account for fatigue from the task. To ensure consistency between the pre-rest and post-rest measurements, careful attention was paid to controlling the posture and environmental conditions. All measurements were conducted in a soundproof laboratory, with room temperature and humidity maintained at comfortable levels.

**Figure 1.**
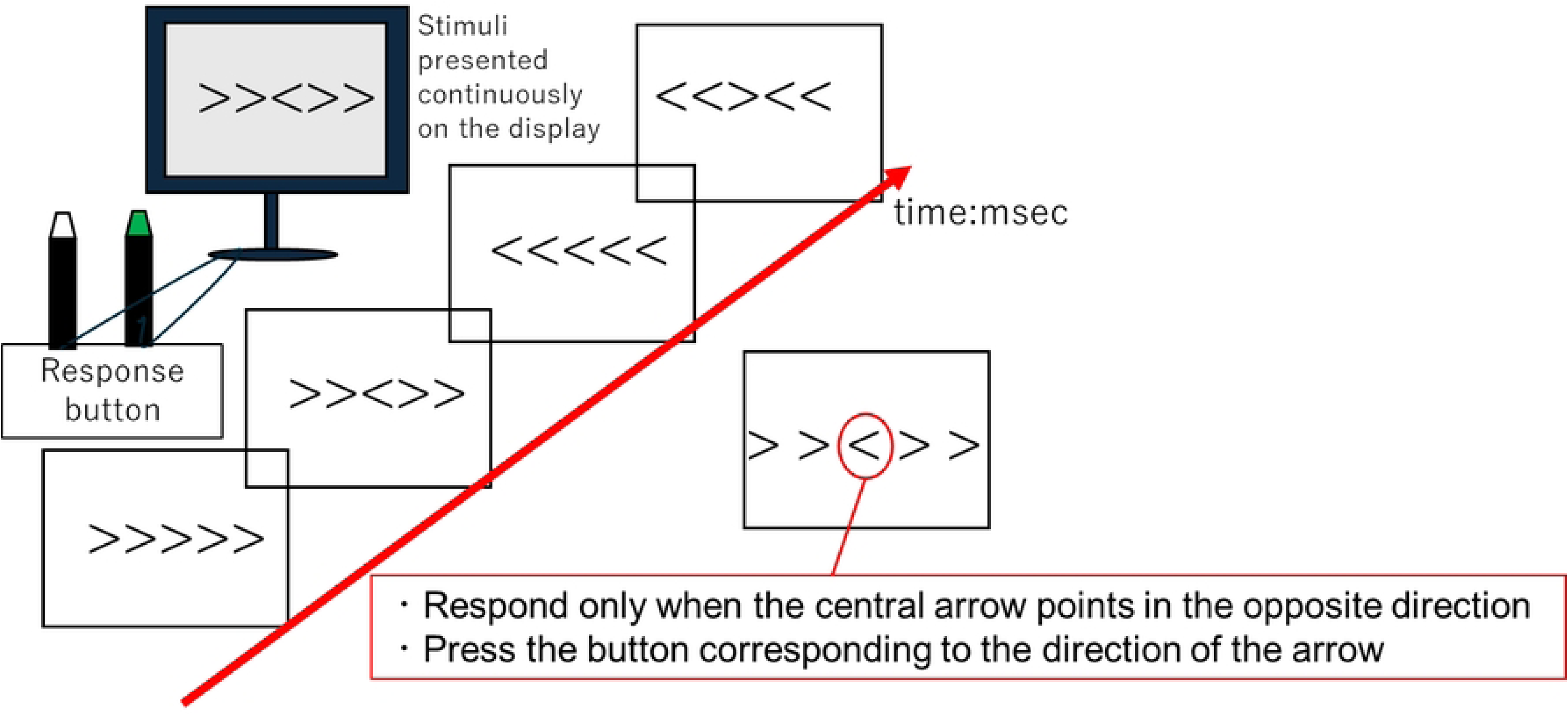
Schematic diagram of the cognitive task system.

### 2.3 Analysis of behavioural indicators

To compare the number of correct responses during the inhibitory task, the number of times each participant correctly responded during the task was recorded. Specifically, the number of times an appropriate button was pressed in response to an incongruent stimulus was counted as a correct response. For the correct response data of each group (young and elderly groups), we first confirmed normality using the Shapiro–Wilk test. To compare the number of correct responses in the inhibition task according to age, we examined the differences in the number of correct responses between the group conditions (elderly and young groups). The significance level for all the tests was set at 5%.

### 2.4 Electroencephalogram measurement and analysis

The measurements were conducted in a shielded room to minimize external electromagnetic noise. For electroencephalogram (EEG) measurements, the bilateral auricles were used as reference electrodes, and 19 scalp electrodes were placed according to the international 10-20 system at the following locations: FP1, FP2, F3, F4, C3, C4, P3, P4, O1, O2, F7, F8, T3, T4, T5, T6, Fz, Cz, and Pz on the scalp based on the international 10-20 method. The sampling frequency was 1000 Hz, and the bandpass filter was set to 0.5–35 Hz. The recorded data were analyzed using BIMUTAS II (Kissei Comtec Co., Ltd.). Responses were classified as correct when the appropriate button was pressed in response to a mismatched stimulus and as incorrect when the wrong button was pressed. Brainwave data spanning 1000 msec, from 300 msec before to 700 msec after the stimulus, were extracted. Additionally, electrooculogram was measured from the eyelid region of the dominant eye, and components > 100 μV were excluded as artifacts. Independent component analysis (ICA) was performed using MATLAB (version R2023b, MathWorks) to process the noise. The latest artifact removal techniques were applied to the preprocessing of the EEG data. Specifically, in addition to ICA, the Artifact Subspace Reconstruction algorithm was used [47]. This enabled a more effective removal of artifacts derived from electromyography and eye movements [48]. The potentials of each component were summed and averaged, and the resulting ERP components were analyzed. Brain activity locations were identified using the LORETA analysis program, a brain functional imaging filter based on coordinate transformation, according to the standard brain based on the Montreal Neurological Institute atlas. In this analysis, we first identified the brain activity regions of ERN and N2 during correct and incorrect responses in the inhibitory task. ERN was analyzed as brain activity during the negative potential onset in the 50–150 msec time window [49]. N2 was analyzed as brain activity during the negative potential onset in the 200–350 msec time window [50]. After identification, data from the 700-msec interval containing all ERP components related to inhibitory function activation were analyzed, and the average activity regions were calculated. To visualize the activity intensities, a color scale based on the current density values was adopted, with regions where the current density values were significantly higher than the 2SD method threshold depicted in red.

Furthermore, to verify the directionality between brain regions showing brain activity during the inhibitory task, we used the “iCoh full vector field” analysis within the LORETA analysis program. For the brain regions identified by LORETA, the brain coordinates of the dorsal and ventral ACC (BA24, 25, 32), left and right dorsolateral prefrontal cortex (DLPFC) (BA9), and left and right FPC (BA10) were defined within a radius of 5 mm from the center coordinates reported in previous studies [51]. Regarding the frequency bands, we defined the 4–7 Hz range as the theta wave band, the 8–13 Hz range as the alpha wave band, and the 14–30 Hz range as the beta wave band.

## 3. Results

### 3.1 Results of behavioral indicators

Statistical analyses were performed using IBM SPSS Statistics for Windows version 27 (IBM). The Shapiro–Wilk test was used to assess data normality. Because the data did not follow a normal distribution, the nonparametric Mann–Whitney U test was adopted. The difference in the number of correct responses in the inhibition task between the elderly and young groups was examined using the Mann–Whitney U test. The results showed that the elderly group had significantly fewer correct responses than the young group (z = –4.99, p < 0.001). The effect size was large (r = 0.88) (Table 1, Fig. 2).

**Figure 2.**
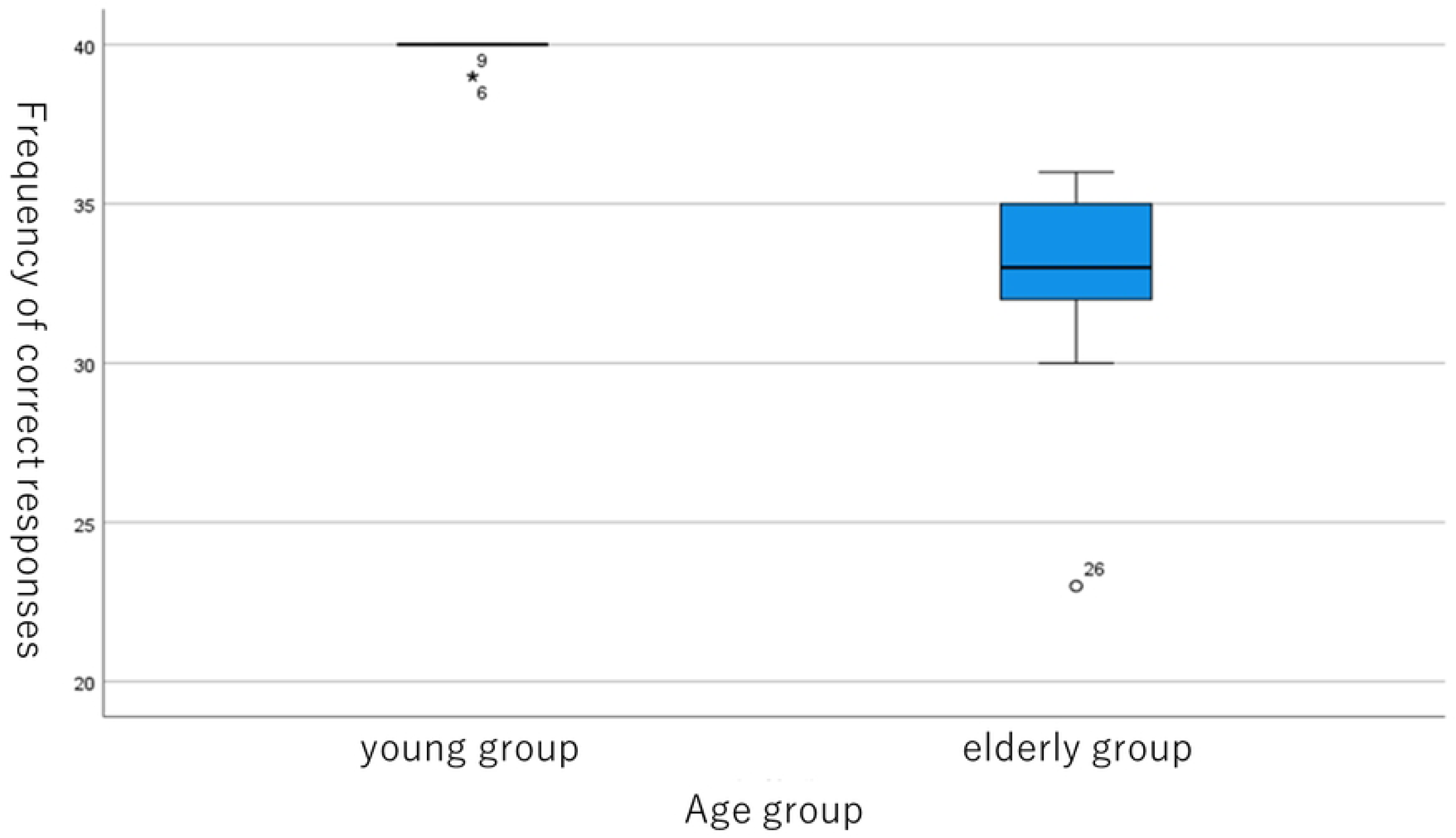
Distribution of correct responses in the inhibitory task according to age group.

**Table 1.**
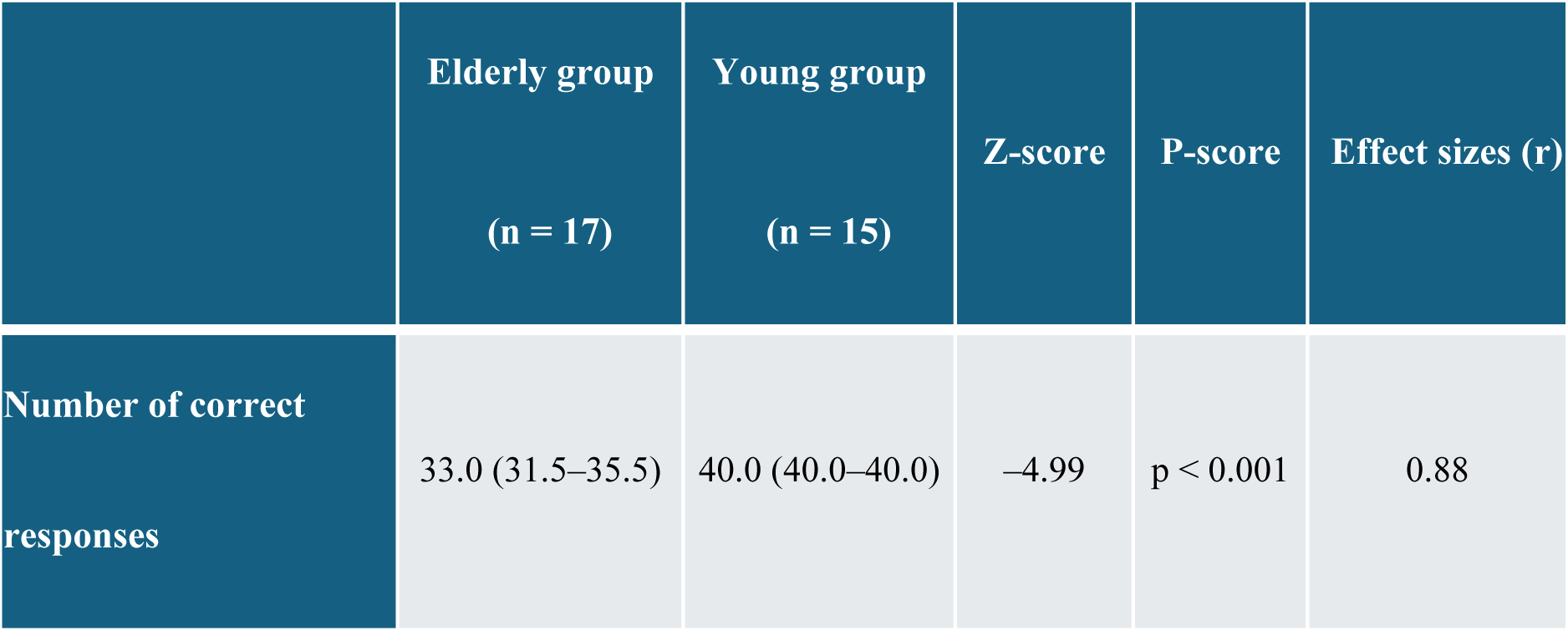
Comparison of correct responses in the inhibitory task between age groups.

|  | Elderly group<br>(n = 17) | Young group<br>(n = 15) | Z-score | P-score | Effect sizes (r) |
| --- | --- | --- | --- | --- | --- |
| Number of correct responses | 33.0 (31.5–35.5) | 40.0 (40.0–40.0) | –4.99 | $p < 0.001$ | 0.88 |

Data are presented as medians (Q1, Q3), where Q1 represents the first quartile (25th percentile) and Q3 represents the third quartile (75th percentile). Group comparisons were performed using the Mann–Whitney U test. Effect sizes were calculated as r and interpreted according to Cohen’s criteria: r = .10 (small), r = .30 (medium), and r = .50 (large).

### 3.2 Brain activity during the inhibitory task in the elderly group

Using LORETA analysis, during the inhibitory task, both during correct and incorrect responses, ERN showed brain activity in the ACC, and N2 showed brain activity in the ACC and FPC. Additionally, during the 700-msec interval associated with inhibitory function, superior brain activity was observed in the ACC, DLPFC, and FPC during both correct and incorrect responses to the inhibitory task (Figures 3 and 4). Results from independent group comparisons using LORETA showed no statistically significant differences between correct and incorrect responses to the inhibitory task.

**Figure 3.**
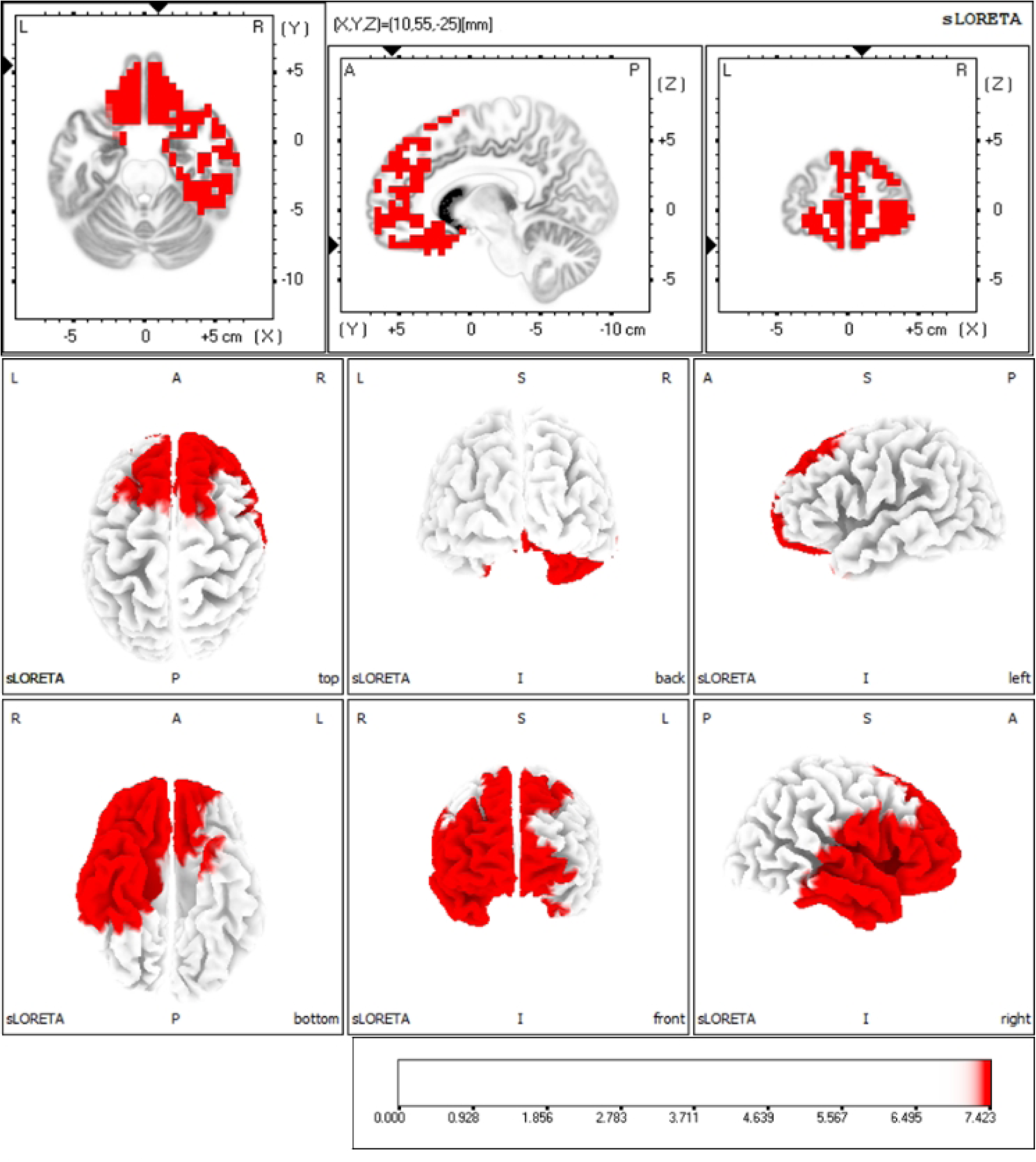
Brain activation regions during correct responses in the inhibitory task in the elderly group.

**Figure 4.**
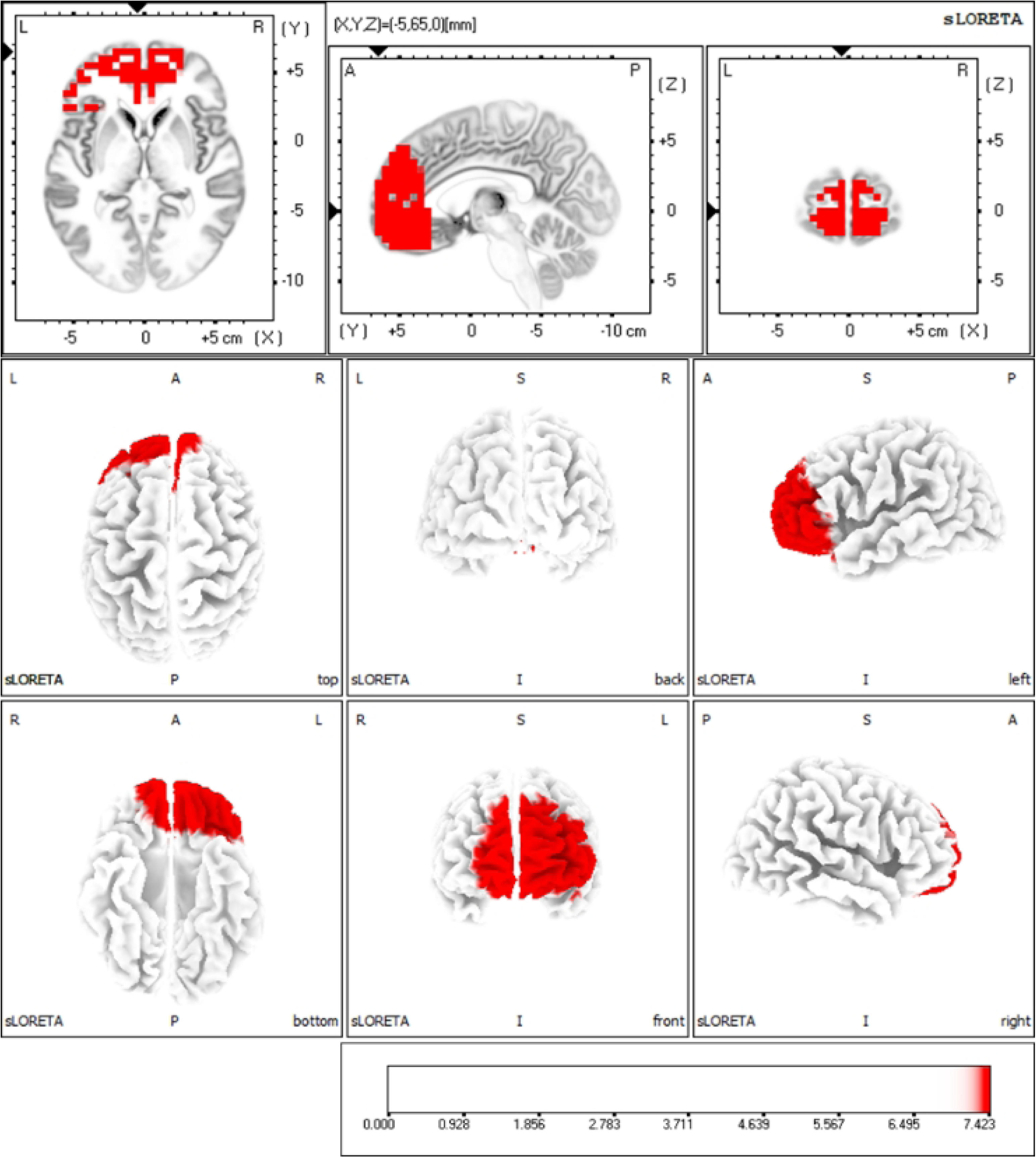
Brain activation regions during incorrect responses in the inhibitory task in the elderly group.

### 3.3 Directionality of brain activity during inhibitory tasks in the elderly group

#### 3.3.1 Functional differences in directionality between inhibitory task correct responses and errors

During inhibitory task correct responses in the elderly group, a significantly stronger directedness was observed from the left DLPFC to the right FPC in the alpha and beta wave bands. Additionally, a significantly stronger directedness was observed from the right FPC to the ventral ACC and both DLPFCs in the alpha and beta wave bands. In contrast, during inhibitory task incorrect responses in the elderly group, a significantly stronger directedness was observed from the ventral ACC to the left FPC in the beta wave band. Additionally, mutually significant stronger directedness was observed between the ventral ACC and dorsal ACC in the theta, alpha, and beta wave bands (Figure 5).

**Figure 5.**
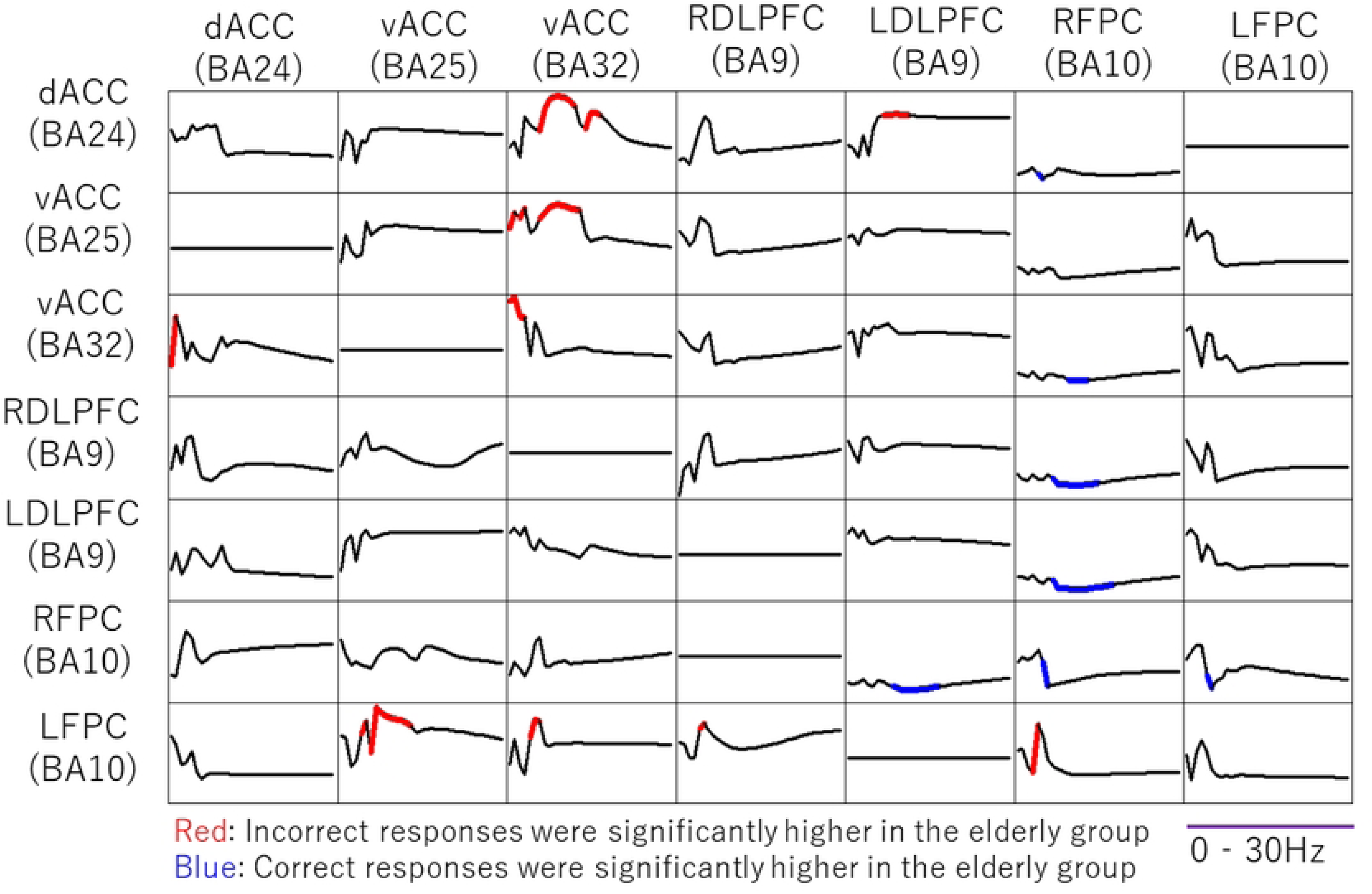
Directed graph comparing incorrect and correct responses in the inhibitory task in the elderly group.

The results of this analysis are presented as a 7 × 7 matrix showing the directional connectivity between brain regions. In each matrix cell, the horizontal axis represents frequency (0–30 Hz), and the vertical axis represents partial directed coherence values (dimensionless). The superimposed red and blue lines indicate statistically significant directional connectivity. The vertical arrangement of the matrix indicates the direction of information flow, with information flowing from the regions displayed horizontally (senders) to the regions displayed vertically (receivers).

### 3.4 Brain activity during the inhibitory task in the young group

During correct responses to the inhibitory task, ERN showed brain activity in the ACC, and N2 showed brain activity in the ACC and FPC. Furthermore, during the 700-msec interval related to inhibitory function, dominant brain activity was observed in the ACC, DLPFC, and FPC during correct responses to the inhibitory task (Figure 6). Directional analysis of correct and incorrect responses to the inhibitory task in the elderly group could not be performed in the young group because the number of data points required for additive averaging in EEG data processing was insufficient, thus preventing the verification of functional differences.

**Figure 6.**
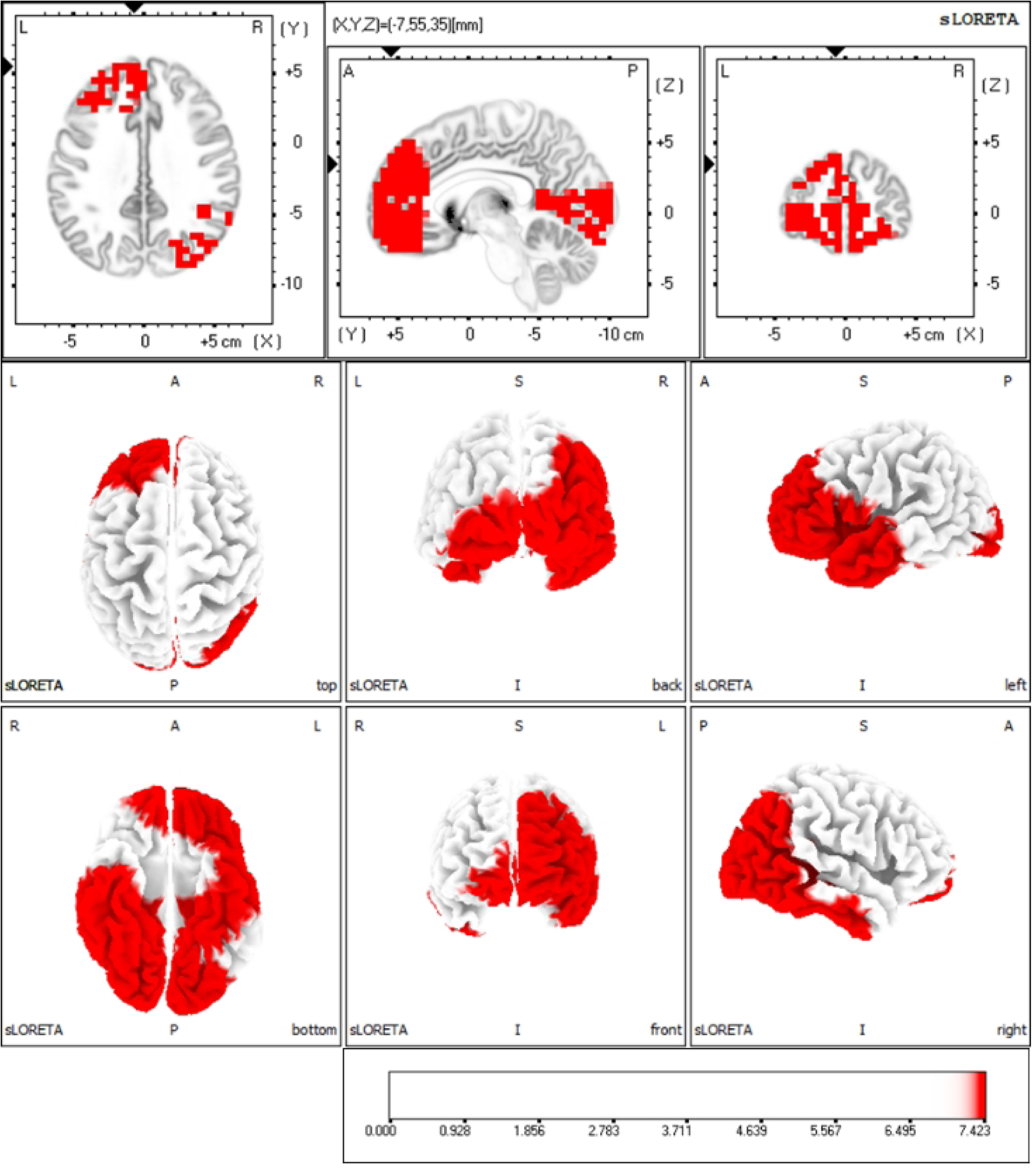
Brain activation regions during correct responses in the inhibitory task in the young group.

### 3.5 Comparison of brain activity between the elderly and young groups

Independent group comparisons of data for the 700-msec interval related to inhibitory function using LORETA revealed that brain activity during correct inhibitory task responses was significantly higher in the young group than in the elderly group in the FPC and occipital lobe.

### 3.6 Comparison of directionality during inhibitory task correct responses between the elderly and young groups

Comparing data from the young group during inhibitory task correct responses with data from the elderly group during inhibitory task correct responses, significantly stronger directionality was observed in the alpha and beta wave bands between the ventral and dorsal ACC in the young group. Additionally, a significantly stronger directionality was observed in the alpha and beta wave bands between the ACC and left and right DLPFC and between the left and right FPC. Furthermore, significant stronger directionality was observed between the DLPFC and FPC in the alpha and beta wave bands (Figure 7).

**Figure 7.**
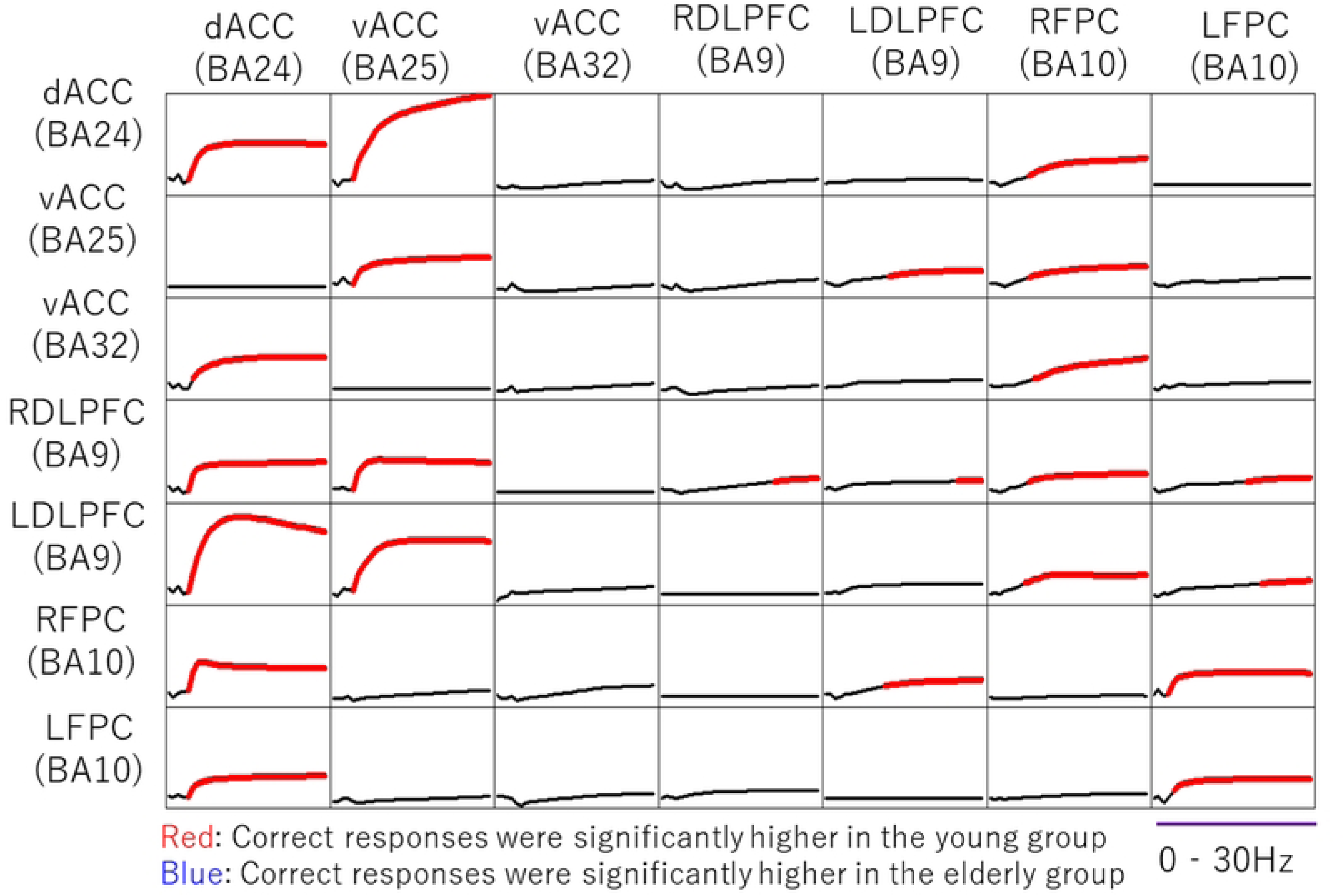
Directed graph comparing correct responses in the inhibitory task between the young and elderly groups.

The results of this analysis are presented as a 7 × 7 matrix showing the directional connectivity between brain regions. In each matrix cell, the horizontal axis represents frequency (0–30 Hz), and the vertical axis represents partial directed coherence values (dimensionless). The superimposed red and blue lines indicate statistically significant directional connectivity. The vertical arrangement of the matrix indicates the direction of information flow, with information flowing from the regions displayed horizontally (senders) to the regions displayed vertically (receivers).

## IV. Discussion

### 4.1 Behavioral indicators

As the Shapiro–Wilk test rejected the assumption of data normality, the Mann–Whitney U test, a nonparametric test, was used to examine the difference in the number of correct responses between the elderly and young groups in the inhibition task. The results showed that the elderly group had significantly fewer correct responses than the young group (z = –4.99, p < 0.001). The effect size was also large (r = 0.88), suggesting a decline in inhibitory function in older adults. A performance decline in older adults evaluating specific executive functions was clearly demonstrated by this task. Verhaeghen et al. [52] reported a meta-analysis examining age-related effects on executive function and attention, suggesting that executive function generally declines with age and that this decline contributes to impaired performance on cognitive tasks. The results of this study support previous findings on age-related changes in cognitive function in older adults. Furthermore, a large-scale meta-analysis published in 2023 demonstrated that inhibitory function decline was more pronounced than that of other executive functions in adults aged ≥ 65 years [53]. Additionally, reports indicate that a decline in inhibitory function is associated with difficulties in activities of daily living [54], which supports the clinical significance of this study.

### 4.2 Neural basis of inhibitory function in the elderly

#### 4.2.1 Brain activity during the appearance of error-related negativity and N2

To examine the neural activity associated with inhibitory function, we compared brainwave data during the 700-msec period following a trigger, focusing on correct and incorrect responses during an inhibitory task. Prior to this analysis, using LORETA analysis, significant brain activity was observed in the ACC and FPC during the appearance of the ERN and N2 components during correct and incorrect responses. These brain regions have previously been reported to be involved in error detection and response inhibition, and the present results are consistent with those of previous studies [55]. These findings indicate that the neural activity associated with inhibitory function is appropriately captured, thereby establishing the validity of the subsequent detailed analyses.

#### 4.2.2 Characteristics of brain activity during the inhibitory task in the correct response condition

Significant directed activity observed in the alpha and beta wave bands from the left DLPFC to the right FPC and from the right FPC to both DLPCs during the correct response in the elderly has been previously reported. These findings suggest that alpha and beta waveband activities in the frontal lobe are closely associated with the control of executive function, maintenance of cognitive flexibility [56], and optimization of performance processes [57]. In particular, the information transmission pathway from the DLPFC to the FPC serves as the foundation for metacognitive processing [58] and enables flexible behavioral control adapted to the situation [59]. Concurrently, reverse information transmission from the FPC to DLPFC contributes to the establishment of effective self-monitoring systems [60]. The bidirectional directionality observed in this study suggests that self-monitoring functions within the metacognitive system operate efficiently and that feedback mechanisms based on information obtained therein are integrated to facilitate performance optimization.

Recent studies indicate a positive relationship between DLPFC–FPC functional connectivity and cognitive flexibility [61]. Additionally, stimulation of the DLPFC using transcranial direct current stimulation improves inhibitory function in older adults [62]. These findings support the importance of the bidirectional connectivity observed between the DLPFC and FPC in this study. Furthermore, regarding the significantly stronger directedness from the right FPC to the ventral ACC in the alpha and beta wave bands, previous studies have reported that activity in the frontal alpha and beta wavebands functions as the neural basis for self-referential information processing [63], the behavioral monitoring system, and metacognitive functions [64]. Furthermore, the FPC plays a central role in metacognitive functions, and the functional connectivity between the ACC and FPC influences the accuracy of metacognitive functions [65]. Based on these findings, the present results suggest that efficient metacognitive functions, such as behavioral monitoring, contribute to the realization of appropriate cognitive control.

These results support the understanding that appropriate functioning of inhibitory control in older adults is dependent on their metacognitive and error detection abilities. Previous studies have also pointed out that the ability to appropriately monitor one’s own cognitive activities and adjust them as needed is essential for maintaining cognitive control in older adults [66]. The findings of this study provide empirical evidence that such interactions and interconnections among cognitive functions can be observed as dynamics within brain networks. Furthermore, in addition to the existing finding that inhibitory function can be improved through cognitive training [67], this study suggests that a comprehensive intervention approach that focuses on enhancing metacognitive abilities is necessary to achieve more effective functional improvements.

#### 4.2.3 Characteristics of brain activity during inhibitory task errors

Compared with correct responses in the elderly group, mutually significant stronger directedness was confirmed in the ACC during error responses across all frequency bands (theta, alpha, and beta). Regarding the network activity within the ACC region, frontal theta waveband activity is involved in unconscious error detection, subsequent cognitive control mechanisms, and emotional response processing [68], whereas frontal alpha waveband activity reflects attention control and the inhibition of unnecessary responses [69]. Additionally, frontal beta waveband activity is closely associated with the fine-tuning of behavior and the processing of feedback information [70]. Furthermore, the ACC itself plays an important role in error detection systems, emotional processing, and the control of inhibitory functions [71]. Specifically, regarding the functional differentiation of the ACC, the dorsal region is responsible for higher-order cognitive control, whereas the ventral region primarily handles emotion-related information processing, suggesting that the ACC as a whole enhances the efficiency of cognitive control [72]. Taken together, these findings suggest that the reciprocal directionality observed within the ACC in this study reflects unconscious error detection and precise inhibitory actions, which play a core role in the behavioral correction process following error responses in inhibitory tasks.

Compared with the correct response condition in the elderly group, significantly stronger directionality from the ventral ACC to the left FPC was observed in the beta wave band during the error response condition. Additionally, compared with the error response condition in the elderly group, significantly stronger directionality from the right FPC to the ventral ACC was observed in the alpha and beta wave bands during the correct response condition. Alpha and beta waveband activities in the frontal lobe are associated with cognitive control functions and higher-order information processing [73] and metacognitive judgment processes [74]. Additionally, regarding the functional connectivity between the ACC and FPC, the FPC functions as a central hub for metacognitive judgment [75], and coordinated activity with the ACC enables the integration of cognitive and emotional information [76]. Based on these findings, the results of this study can be interpreted as reflecting a process in which negative feedback information generated by error responses is transmitted from the ventral ACC to the left FPC, thereby promoting the higher-order processing of that information. Furthermore, activation of metacognitive judgment functions in the right FPC suggests that it contributes to the realization of appropriate cognitive control.

These findings provide important insights into the role of ACC activity in the error correction process in older adults, suggesting that ACC activity plays a central role and is finely tuned by input from the FPC. Previous studies have also suggested that the functional connectivity between the ACC and FPC plays an important role in cognitive control in older adults [77]. In particular, the FPC is a brain region less affected by age-related structural and functional changes [78] and is considered to play a crucial role in maintaining cognitive function in older adults. The results of this study suggest that these functions of the FPC are exerted through coordinated activity with the ACC.

### 4.3 Age-related changes in cognitive function

#### 4.3.1 Age-related changes in error detection ability and their association with inhibitory function

In a comparison between the older and younger groups, mutual directionality between the ventral ACC and dorsal ACC was observed only in the younger group in the alpha and beta wave bands, which is considered to reflect age-related changes in error detection ability and inhibitory function. The ACC is involved in behavioral monitoring and detection of cognitive conflicts, playing a crucial role as the neural basis for error detection ability [79]. Specifically, the dorsal ACC is involved in the execution of cognitive control, whereas the ventral ACC is associated with processing emotional responses [80]. These regions interact to efficiently process information, enabling appropriate cognitive activities, such as error detection. The reciprocal directed interactions observed within the ACC in the young group suggest that information processing related to error detection is being efficiently performed. However, the absence of such interactions within the ACC in the elderly group suggests a decline in error detection ability with age. This suggests that an age-related decline in error detection ability may be closely related to a decline in inhibitory function. Age-related declines in error detection ability and inhibitory function occur simultaneously [81]. The ACC utilizes the information generated by error detection to adjust the inhibitory function. Therefore, ACC dysfunction may affect both error detection ability and inhibitory function. The results of this study provide insights into age-related functional changes in the ACC from the perspective of brain network dynamics.

#### 4.3.2 Age-related changes in immediate cognitive control and their association with inhibitory function

In comparisons between the elderly and young groups, the significant stronger directedness observed in the alpha and beta wave bands between the ACC and DLPFC in the young group has been reported in previous studies to indicate that the ACC is involved in detecting cognitive competition [82] and that this information promotes immediate inhibitory behavioral control in the DLPFC [83]. Furthermore, evidence suggests that the ACC and DLPFC achieve efficient cognitive control through parallel information processing and integration [84].

Considering these reports, the results of this study are thought to reflect an immediate inhibitory behavioral control mechanism mediated by a coordinated and parallel information processing system between the ACC and DLPFC. This suggests that self-referential information processing and behavioral monitoring within the metacognitive system play essential roles in achieving appropriate cognitive control and immediate inhibitory behavioral control, providing evidence that these functions decline with age.

### 4.4. Interaction mechanisms among inhibitory function, error detection ability, and metacognitive ability

The results of this study suggest that the interaction mechanism among inhibitory function, unconscious error detection ability, and metacognitive ability involves the functional integration of the ACC and FPC, which plays a crucial role in the integration of these functions. The ACC is involved in inhibitory control and unconscious error detection, whereas the FPC is associated with metacognitive monitoring. Coordinated activity between these regions contributes to the realization of appropriate cognitive control. Second, this study suggests that age-related declines in inhibitory function may be associated with reductions in unconscious error detection and metacognitive abilities.

Comparing the elderly and young groups, this study found that the young group showed superior functional connectivity between the ACC and FPC, suggesting that age-related declines in metacognitive ability may contribute to the decline in inhibitory function. The decline in cognitive function with age is associated with the interaction between multiple cognitive functions [85]. Metacognitive ability is a higher-order function that regulates the efficiency of other cognitive functions, and its decline leas to a decline in other cognitive functions [86]. The results of this study empirically demonstrate the role of metacognitive ability in relation to inhibitory function and error detection ability. Further investigations are necessary to determine the effects of interventions aimed at improving error detection and metacognitive abilities on the maintenance and improvement of other cognitive functions, including inhibitory functions.

## V. Limitations and future prospects of the present study

One limitation of this study is its relatively small sample size. In particular, the number of participants in the elderly group was limited to 17; therefore, caution should be exercised when generalizing the results. From the perspective of statistical power, verification using a larger sample size is desirable. Furthermore, this study only used the flanker task as an inhibitory task. However, we consider it important to examine the generalizability of the findings regarding correlations with other types of inhibitory tasks, such as the Stroop and Go/No-go tasks. Additionally, more detailed consideration is required regarding the difficulty level of the tasks. This study focused on comparisons between age groups; thus, it is possible that individual differences, such as educational background and cognitive ability, may have influenced the results. These factors affect the cognitive function and brain activity [87]. Educational background is associated with cognitive reserve and may have a protective effect against age-related changes in cognitive function [88]. The extent of daily intellectual and social activities should also be considered a factor that may have influenced our results. Future studies should examine these factors while appropriately controlling for them. Furthermore, because this study adopted a cross-sectional design, it was not possible to directly examine age-related changes. A longitudinal study design that tracks changes within individuals could allow for a more detailed understanding of age-related changes in cognitive function and brain activity [89]. In particular, as cognitive function exhibits significant individual differences, a longitudinal approach can help clarify the factors underlying these individual differences. Furthermore, although this study focused only on healthy elderly individuals, including those with MCI or early-stage dementia, it provides a more comprehensive understanding of the mechanisms underlying cognitive decline [90]. Particularly, the relationship between inhibitory function and changes in metacognitive ability during cognitive decline is an important research topic.

The findings of this study provide important insights for the early detection of cognitive decline and development of preventive interventions. In particular, the establishment of noninvasive evaluation methods using EEG is expected to enhance the feasibility of cognitive function screening at the community level. It is anticipated that machine learning algorithms can be used to construct predictive models of cognitive decline risk based on the neural network characteristics identified in this study [91]. Additionally, the development and validation of intervention programs aimed at improving metacognitive abilities and error detection abilities are important challenges [92]. Furthermore, it is worth investigating the potential of non-invasive brain stimulation methods, such as transcranial electrical stimulation, for intervention [93]. By integrating these study findings, a comprehensive approach for the prevention and early intervention in cognitive decline can be established.

## Author Contributions

Conceptualization, K.U. and T.K.; data curation, K.U., K.N. (Nishimoto Kazuhei), H.I., R.Y., and K.M. (Maeda Kouta); formal analysis, K.U., K.N. (Nishimoto Kazuhei), O.K., S.M., K.M. (Morita Kiichiro), and T.K.; methodology, K.U., S.M., K.M. (Morita Kiichiro), and T.K.; supervision, T.K.; validation, K.U., K.N. (Nishimoto Kazuhei), H.I., R.Y., K.M. (Maeda Kouta), and T.K.; visualization, K.U.; writing – original draft, K.U., K.N. (Nishimoto Kazuhei), and T.K.; writing – review & editing, K.U. and T.K.

## Institutional Review Board Statement

This study was conducted in accordance with the Declaration of Helsinki and approved by the Institutional Review Board of Kyoto Tachibana University (protocol code 22–03, April 22, 2022).

## Informed Consent

Informed consent was obtained from all the participants involved in this study.

## Data Availability

The datasets generated and/or analyzed during the current study are not publicly available because they contain information that can compromise study participants’ privacy/consent but are available from the corresponding author upon reasonable request.

## Abbreviations

The following abbreviations are used in this manuscript:

ACC: Anterior Cingulate Cortex
DLPFC: Dorsolateral Prefrontal Cortex
EEG: Electroencephalography
ERN: Error-Related Negativity
ERP: Event-Related Potentials
FPC: Frontal Polar Cortex
ICA: Independent Component Analysis
LORETA: Low-Resolution Electromagnetic Tomography
MCI: Mild Cognitive Impairment

## References

1. Cabeza R, Albert M, Belleville S, et al. Maintenance, reserve and compensation: the cognitive neuroscience of healthy ageing. Nat Rev Neurosci. 2024;25(2):98–115.

2. Hedden T, Gabrieli JD. Insights into the ageing mind: a view from cognitive neuroscience. Nat Rev Neurosci. 2023;24(8):482–496.

3. Damoiseaux JS. Effects of aging on functional and structural brain connectivity. NeuroImage. 2023;260:119496.

4. Andrews-Hanna JR, Snyder AZ, Vincent JL, et al. Disruption of large-scale brain systems in advanced aging. Neuron. 2024;112(3):458–471.

5. Salthouse TA. Trajectories of normal cognitive aging. Psychol Aging. 2023;38(1):45–58.

6. Stern Y. What is cognitive reserve? Theory and research application of the reserve concept. Journal of the International Neuropsychological Society. 2002;8(3):448–460.

7. Cabeza R, Dennis NA. Frontal lobes and aging: Deterioration and compensation. In: Principles of Frontal Lobe Function. 2023:628-652.

8. Tuck er-Drob EM. Cognitive aging and dementia: A life-span perspective. Annu Rev Dev Psychol. 2023;5:177–196.

9. Livingston G, Huntley J, Sommerlad A, et al. Dementia prevention, intervention, and care: 2024 report of the Lancet Commission. Lancet. 2024;403(10452):131–178.

10. Jack CR Jr, Bennett DA, Blennow K, et al. NIA-AA Research Framework: Toward a biological definition of Alzheimer’s disease. Alzheimers Dement. 2024;20(1):47–94.

11. Sperling RA, Aisen PS, Beckett LA, et al. Toward defining the preclinical stages of Alzheimer’s disease: recommendations from the National Institute on Aging-Alzheimer’s Association workgroups. Alzheimers Dement. 2011;7(3):280–292.

12. Petersen RC, Lopez O, Armstrong MJ, et al. Practice guideline update summary: Mild cognitive impairment. Neurology. 2023;100(3):126–135.

13. Buckley RF, Hanseeuw B, Schultz AP, et al. Functional brain changes in preclinical-–Alzheimer’s disease. Nat Neurosci. 2024;27(2):234–245.

14. Maass A, Berron D, Harrison TM, et al. Alzheimer’s pathology targets distinct memory networks in the ageing brain. Brain. 2023;146(4):1456–1470.

15. Rodrigues EA, Moreno PI, Gutierrez-Ruiz K, et al. Machine learning approaches for early detection of cognitive decline using EEG biomarkers. Clin Neurophysiol. 2024;155:88–102.

16. Sherman DS, Mauser J, Boots EA, et al. The efficacy of cognitive intervention in mild cognitive impairment. JAMA Neurol. 2023;80(5):470–478.

17. Teunissen CE, Verberk IMW, Thijssen EH, et al. Blood-based biomarkers for Alzheimer’s disease. Nat Med. 2022;28(5):910–921.

18. Fjell AM, Walhovd KB. Structural brain changes in aging: courses, causes and cognitive consequences. Rev Neurosci. 2023;34(8):879–904.

19. Miyake A, Friedman NP, Emerson MJ, et al. The unity and diversity of executive functions and their contributions to complex “Frontal Lobe” tasks. Cognitive Psychology. 2000;41(1):49–100.

20. Miyake A, Friedman NP. The nature and organisation of individual differences in executive functions. Current Directions in Psychological Science. 2022;31(1):8–14.

21. Aron AR, Robbins TW, Poldrack RA. Inhibition and the right inferior frontal cortex. Trends Cogn Sci. 2004;8(4):170–177.

22. Cai W, Ryali S, Chen T, et al. Dynamic interactions within the frontoparietal network during inhibitory control. Cereb Cortex. 2024;34(2):456–469.

23. Friedman NP, Robbins TW. The role of prefrontal cortex in cognitive control and executive function. Neuropsychopharmacology. 2022;47(1):72–89.

24. Aron AR, Robbins TW, Poldrack RA. Right inferior frontal cortex: addressing the rebuttals. Front Hum Neurosci. 2023;17:1145797.

25. Zelazo PD, Carlson SM. The neurodevelopment of executive function skills. Journal of Child Psychiatry and Psychology. 2024;65(2):156–179.

26. Rey-Mermet A, Gade M. Inhibition in aging: A meta-analysis of Stroop and stop-–signal tasks. Psychol Aging. 2024;39(1):78–92.

27. Miller EK, Lundqvist M, Bastos AM. Working memory 2.0. Neuron. 2023;111(3):345–359.

28. Menon V, D’Esposito M. The role of PFC networks in cognitive control. Annu Rev Neurosci. 2024;47:234–256.

29. Baddeley A. Working memory: looking back and looking forward. Nat Rev Neurosci. 2003;4(10):829–839.

30. Clayton MS, Yeung N, Cohen Kadosh R. The roles of cortical oscillations in sustained attention. Trends Cogn Sci. 2023;27(5):456–468.

31. Falkenstein M, Hohnsbein J, Hoormann J, Blanke L. Effects of crossmodal divided attention on late ERP components. II. Error processing in choice reaction tasks. Electroencephalogr Clin Neurophysiol. 1991;78(6):447–455.

32. Wessel JR. An adaptive theory of error processing. Psychophysiology. 2023;60(3):e14234.

33. Fleming SM, Daw ND. Self-evaluation of decision-making: A general Bayesian framework for metacognitive computation. Psychol Rev. 2023;130(1):45–67.

34. Mansouri FA, Koechlin E, Rosa MGP, Buckley MJ. Managing competing goals — a key role for the frontopolar cortex. Nat Rev Neurosci. 2023;24(11):678–692.

35. Vaccaro AG, Fleming SM. Thinking about thinking: A coordinate-based meta-analysis of neuroimaging studies of metacognitive judgements. Brain Neurosci Adv. 2023;7:123–145.

36. Rouault M, McWilliams A, Allen MG, Fleming SM. Prefrontal mechanisms of metacognitive control. Science. 2023;381(6654):234–239.

37. Bush G, Luu P, Posner MI. Cognitive and emotional influences in the anterior cingulate cortex. Trends Cogn Sci. 2000;4(6):215–222.

38. Holroyd CB, Verguts T. The best laid plans: Computational principles of ACC. Trends Cogn Sci. 2023;27(4):345–356.

39. Gehring WJ, Goss B, Coles MG, et al. A neural system for error detection and compensation. Psychol Sci. 1993;4(6):385–390.

40. Nieuwenhuis S, Ridderinkhof KR, Blom J, et al. Error-related brain potentials are differentially related to awareness of errors. Journal of Neuroscience. 2023;43(12):2345–2356.

41. Botvinick MM, Braver T. Motivation and cognitive control: From behaviour to neural mechanisms. Annu Rev Psychol. 2023;74:345–367.

42. Ridderinkhof KR, Ullsperger M, Crone EA, Nieuwenhuis S. Neurocognitive mechanisms of cognitive control. Nat Rev Neurosci. 2024;25(2):123–139.

43. Alexander WH, Brown JW. Computational models of ACC. Current Opinion in Behavioural Sciences. 2024;55:89–96.

44. Shenhav A, Botvinick MM, Cohen JD. The expected value of control: An integrative theory of ACC. Neuron. 2023;111(4):567–582.

45. Cole MW, Ito T, Schultz D, et al. The frontoparietal control system. Annu Rev Neurosci. 2023;46:567–589.

46. Folstein MF, Folstein SE, McHugh PR. “Mini-mental state”: A practical method for grading the cognitive state of patients for the clinician. J Psychiatr Res. 1975;12(3):189–198.

47. Chang CY, Hsu SH, Pion-Tonachini L, Jung TP. Evaluation of artifact subspace reconstruction for automatic EEG artifact removal. IEEE Transactions on Neural Systems and Rehabilitation Engineering. 2023;31:234–245.

48. Pion-Tonachini L, Kreutz-Delgado K, Makeig S. ICLabel: An automated electroencephalographic independent component classifier. NeuroImage. 2023;258:119359.

49. Cavanagh JF, Frank MJ. Frontal theta as a mechanism for cognitive control. Trends Cogn Sci. 2024;28(1):67–79.

50. Folstein JR, Van Petten C. After the P3: Late executive processes in stimulus categorisation. Psychophysiology. 2023;60(2):e14189.

51. Badre D, Nee DE. Frontal cortex and the hierarchical control of behaviour. Trends Cogn Sci. 2023;27(2):156–169.

52. Verhaeghen P, Cerella J. Aging, executive control, and attention: a review of meta-analyses. Neuroscience & Biobehavioral Reviews. 2002;26(7):849–857.

53. Harada CN, Natelson Love MC, Triebel KL. Normal cognitive aging and activities of daily living. Clin Geriatr Med. 2023;39(1):123–136.

54. Zanto TP, Gazzaley A. Aging of the frontal lobe network. Trends Cogn Sci. 2024;28(3):234–248.

55. Cooper PS, Darriba Á, Karayanidis F, Barceló F. Frontal theta predicts specific cognitive control-induced behavioural changes. NeuroImage. 2023;265:119–132.

56. Schmidt R, Ruiz MH, Kilavik BE, et al. Beta oscillations in cognitive control. Annu Rev Psychol. 2023;74:567–589.

57. Koechlin E. Prefrontal executive function and adaptive behaviour. Current Opinion in Behavioural Sciences. 2024;55:101–108.

58. Dixon ML, De La Vega A, Mills C, et al. Heterogeneity within the frontoparietal control network. Cereb Cortex. 2023;33(5):2345–2358.

59. Seeley WW. The salience network: A neural system for perceiving salience. Annu Rev Neurosci. 2023;46:345–367.

60. Fleck MS, Daselaar SM, Dobbins IG, Cabeza R. Metacognitive awareness in aging. Psychol Aging. 2024;39(2):234–248.

61. Hsu WY, Zanto TP, Anguera JA, et al. Effects of transcranial direct current stimulation on cognitive function in healthy older adults: A meta-analysis. Brain Stimulation. 2023;16(2):456–467.

62. Hampshire A, Sharp DJ. Contrasting network and modular perspectives on inhibitory control. Trends Cogn Sci. 2023;27(6):567–579.

63. Burgess PW, Dumontheil I, Gilbert SJ. The gateway hypothesis of rostral prefrontal cortex function. Trends Cogn Sci. 2023;27(8):678–691.

64. Euston DR, Gruber AJ, McNaughton BL. The role of medial prefrontal cortex in memory and decision making. Neuron. 2023;111(7):1234–1248.

65. Hertzog C, Dunlosky J. Metacognition in later adulthood. Current Directions in Psychological Science. 2023;32(3):234–241.

66. Simons DJ, Boot WR, Charness N, et al. Do “brain-training” programmes work? Psychol Sci Public Interest. 2023;24(1):45–89.

67. Cohen MX. A neural microcircuit for cognitive conflict detection and signalling. Trends Neurosci. 2024;47(2):123–135.

68. Vassena E, Holroyd CB, Alexander WH. Computational psychiatry of cognitive control. Biol Psychiatry. 2023;93(8):678–689.

69. Silvetti M, Nuñez Castellar E, Roger C, Verguts T. ACC as a model-based reinforcement learning system. Psychol Rev. 2024;131(1):123–145.

70. Botvinick MM, Cohen JD, Carter CS. Conflict monitoring and anterior cingulate cortex: an update. Trends Cogn Sci. 2004;8(12):539–546.

71. Carter CS, van Veen V. ACC and conflict detection: An update. Cogn Affect Behav Neurosci. 2023;23(3):456–468.

72. MacDonald AW 3rd, Cohen JD, Stenger VA, Carter CS. Specificity of prefrontal dysfunction. American Journal of Psychiatry. 2024;181(2):234–245.

73. Koechlin E, Hyafil A. Anterior prefrontal function and the limits of human decision-making. Science. 2023;382(6668):326–329.

74. Fleming SM, Dolan RJ. The neural basis of metacognitive ability. Philos Trans R Soc Lond B Biol Sci. 2023;378(1885):20210497.

75. Rouault M, Fleming SM. Formation of global self-beliefs in the brain. Proc Natl Acad Sci USA. 2023;120(37):e2306153120.

76. Dosenbach NUF, Fair DA, Miezin FM, et al. Distinct brain networks for adaptive control. Proc Natl Acad Sci USA. 2023;120(25):e2303318120.

77. Campbell KL, Schacter DL. Aging and the resting state: Cognition is not at rest. Current Opinion in Behavioural Sciences. 2023;52:101287.

78. Nyberg L, Lindenberger U. Brain maintenance and cognition in old age. Annu Rev Psychol. 2024;75:234–256.

79. Ridderinkhof KR, Ullsperger M, Crone EA, Nieuwenhuis S. The role of the medial pre frontal cortex in cognitive control. Science. 2004;306(5695):443–447.

80. Devinsky O, Morrell MJ, Vogt BA. Contributions of the anterior cingulate cortex to behaviour. Brain. 1995;118(Pt 1):279–306.

81. Larson MJ, Clayson PE. The relationship between cognitive performance and electrophysiological indices of performance monitoring. Cognitive and Affective Neuroscience. 2023;23(2):234–250.

82. Kerns JG, Cohen JD, MacDonald AW 3rd, et al. Anterior cingulate conflict monitoring and adjustments in control. Science. 2004;303(5660):1023–1026.

83. Miller EK, Cohen JD. An integrative theory of prefrontal cortex function. Annu Rev Neurosci. 2001;24:167–202.

84. Egner T. Principles of cognitive control over task focus and task switching. Nat Rev Psychol. 2023;2(7):405–420.

85. Park DC, Reuter-Lorenz P. The adaptive brain: Aging and neurocognitive scaffolding. Annu Rev Psychol. 2023;74:789–812.

86. Dunlosky J, Hertzog C. Metacognitive judgments and their accuracy in older adults. Psychol Aging. 2023;38(5):567–580.

87. Stern Y, Barnes CA, Grady C, et al. Cognitive reserve in ageing. Lancet Neurol. 2023;22(4):345–356.

88. Valenzuela MJ, Sachdev P. Brain reserve and cognitive decline. Int Psychogeriatr. 2023;35(8):423–435.

89. Lindenberger U. Human cognitive aging: Correcting the fortune? Science. 2024;383(6685):766–770.

90. Albert MS, DeKosky ST, Dickson D, et al. The diagnosis of mild cognitive impairment. Alzheimer’s Dement. 2023;19(9):3841–3852.

91. Parra MA, Butler S, McGeown WJ, et al. Machine learning for dementia prediction. Alzheimer’s & Dementia. 2023;19(5):2014–2027.

92. Rebok GW, Ball K, Guey LT, et al. Ten-year effects of cognitive training. J Am Geriatr Soc. 2024;72(1):96–106.

93. Birba A, Ibáñez A, Sedeño L, et al. Non-invasive brain stimulation in dementia. Annals of Neurology. 2023;94(5):856–872.

